# Predicting Near-term Mortality in Heart Failure: External Validation of Electronic Health Record-Based Deep Learning Model

**DOI:** 10.1101/2025.08.13.25333636

**Authors:** Martha M.O. McGilvray, Amit A. Pawale, Sophia R. Pyeatte, Hailey M. Shepherd, Adam Wilcox, Jeffrey T. Heaton, Michael K. Pasque

**Affiliations:** School of Medicine, Washington University in St. Louis, St. Louis, Missouri; Sever Institute, McKelvey School of Engineering, Washington University in St. Louis, St. Louis, Missouri

**Keywords:** Heart failure, electronic health record (EHR), outcome prediction model, machine learning, deep learning, deep neural network, mechanical circulatory support, cardiac transplantation

## Abstract

**Background:** The dire consequences of heart failure (HF) patient non-response to guideline directed medical therapy often fuel early, non-selective referral for surgical intervention (ventricular assist device [VAD] or transplant). The high-risk associated with these interventions mandates precision in directing them only toward those patients who would otherwise suffer severe near-term deterioration. We previously reported a 52,265-patient deep learning model that predicted 1-year severe decompensation/death in HF inpatients, with a C-statistic of 0.91. We now present external model validation. Few groups applying deep learning to large-scale datasets have achieved external validation using equally large-scale independent datasets, yet proof of generalization is essential to practical applicability.

**Methods:** Our previous study used standard electronic health record (EHR) data to build ensemble deep learning models employing time-series and densely connected networks. The positive-class included both all-cause mortality and referral for HF surgical intervention within 1 year. In the current study, we assessed generalization of model architecture in an external validation test set from the Veterans Cardiac Health and Artificial Intelligence Model Predictions (V-CHAMPS) challenge, a synthetic national governmental sample using a distinct EHR system. While V-CHAMPS is a robust dataset, variables that capture VAD/transplant referral were not readily extracted, limiting the positive-class to mortality only.

**Results:** A total of 380,441 distinct admissions from 75,086 HF patients contributed >720 million EHR datapoints. 23% of observations fit positive-class criteria. The model C-statistic in the external-validation cohort was 0.79.

**Conclusions:** Despite being developed in a single-center dataset with a more precise positive-class, our model architecture maintained relative accuracy when applied to a national sample in an unrelated EHR system. This supports clinical relevancy of the deep-learning model and adaptability with retraining to disparate contexts. This broad applicability suggests considerable potential of EHR-based deep learning models to assist HF clinicians in improving the usage of advanced surgical therapy.

## Introduction

Heart failure (HF) patients comprise the largest and most expensive cardiovascular disease subgroup in the United States (US), a subgroup that continues to increase in both size and impact [4]. Current overall annual US HF mortality approaches 22-30% [2,3]. The sickest class of these patients, those in New York Heart Association (NYHA) class IV, have an annual mortality greater than 50% [1].

Alarming congestive symptoms and severely impaired left ventricular contractile function often characterize initial HF patient presentation. Despite the gravity of their presentation, most HF patients subsequently demonstrate a favorable response to guideline directed medical therapy (GDMT) [5–8]. However, the minority who do not respond well to GDMT will often precipitously decline, experiencing severe end-organ injury or death, precluding any salvage attempt with surgical intervention [9]. Patients who undergo surgical therapy when it would not ultimately have been needed face undue high procedural risk and a significant impact to their quality of daily life, particularly with VAD implantation [9–11]. There is a fine and difficult-to-achieve balance in deciding which patients to refer for early surgical management of HF, with potentially dire consequences for errors in judgement. Despite this, current standard-of-care metrics used to make these decisions (e.g. left ventricular ejection fraction [LVEF], blood urea nitrogen [BUN], serum sodium), perform far below the level of generalizable accuracy needed to truly help clinicians make accurate decisions in what therapy to offer their patients [5,12–15]. This area therefore presents the perfect use-case for deployment of deep learning outcome-predictive models.

To address the clinical need for more accurate patient-specific, near-term outcome prediction in HF patients, we previously developed an electronic health record (EHR)- based predictive deep learning model in a large, single-center cohort of 52,265 HF patients. This recurrent neural network (RNN)-based model, previously reported in this journal [16], used only standard EHR variables to predict 1-year HF death or severe decompensation (ventricular assist device [VAD] or transplant) with a predictive area under the receiver operating characteristic (ROC) curve (AUC) of 0.91. Prior to prospective clinical application, confirmation of accuracy in a separate external validation test set of HF patients not used in original model training is necessary. The ideal test set to assess model generalizability across the US HF population is a truly national sample, i.e. one that does not simply draw patients from a few selected states or centers.

Although few such national samples are readily accessible, the Veterans Health Association (VHA) Veterans Cardiac Health and AI Model Predictions (V-CHAMPS) HF challenge provides a large synthetic data lake well-suited for this objective. The VHA V-CHAMPS HR data lake is not only large, robust, and well-documented, but is also comprised of patients from every regional subgroup in our target US HF population.

This synthetic dataset was generated to reflect real patient data while preserving privacy, enabling model validation without compromising protected health information.These attributes make it an ideal follow-up validation test set for our original 52,265-patient-based 1-year HF outcome predictive model to determine whether our model generalizes to data obtained from sources beyond a single institution.

Although similar HF outcome predictive machine learning models have been developed, few have been subjected to subsequent rigorous clinical validation in an external validation test set unrelated to the model’s original training data. Successful external validation confirms clinical applicability of the model and offers real-world estimation of model generalization.

## Methods

### Data Source

This study was approved by the Washington University School of Medicine Human Studies Institutional Review Board. Our original predictive model was developed from EHR variables derived from a decade of individual HF patient records at a single large academic institution, Barnes-Jewish Hospital at Washington University Medical Center in St. Louis, Missouri. Of note, the positive class in this model included not only 1-year all-cause mortality, but also referral for placement of mechanical circulatory support (MCS; i.e. VAD or extracorporeal membrane oxygenation [ECMO]) and/or cardiac transplantation.

The large EHR dataset employed in the current investigation for the validation of our original 1-year predictive model was made available by access to the VA V-CHAMPS HF patient data lake [17]. The VHA V-CHAMPS HR patient data lake was created using MDClone to generate non-reversible artificial patient data using the original VA patient records as input. The underlying structure of the original patient data – that is statistical properties and complex relationships between individual data attributes – are preserved in a manner that ensures reversing back to the original dataset is not possible. The resulting artificial VHA data provided an EHR-based validation test set with a well-documented positive class defined by 1-year all-cause mortality.

### Cohort and Study Design

All adult patients included in the complete VHA V-CHAMPS HF dataset comprised our initial validation study cohort (n=133,252 patients). After application of exclusion criteria, our final validation dataset included all the 75,086 patients who fit our study age criteria (≥ 18 < 90 years of age) and had complete patient-specific values in each of the V-CHAMPS datasets (thereby excluding 58,116 patients who had incomplete datasets). The included V-CHAMPS datasets were *lab_results_train.csv*, *demographics_static_train.csv*, *measurements_blood_pressure_train.csv*, *measurements_train.csv*, *medications_administered_train.csv*, *conditions_train.csv*, *procedures_train.csv, inpatient_admissions_train.csv,* and *death_train.csv*.

Our deep learning approach addresses the outcome predictive impact of the chronicity of HF with the goal of providing HF clinicians with near-term outcome predictive power at any point in the clinical course. Practically speaking, clinician need for accurate patient-specific HF outcome prediction is usually hospitalization focused, as is the accumulation of EHR data to support meaningful model development. Although each VHA HF patient contributed at least one hospital admission as a model observation, our inclusion of all available hospital admissions that met study criteria as model observations allowed each patient to contribute more than one observation (if available).

Current HF management paradigms readily acknowledge the potential predictive power of the trends and patterns routinely present in EHR data over the course of chronic HF, including numerous prior outpatient visits and hospitalizations. Accordingly, EHR data from up to a maximum of 100 prior outpatient visits or inpatient hospitalizations that occurred prior to each of the observation hospital admissions were also included in each admission-based observation.

An observation-specific 1-year clock, against which all-cause mortality was referenced for determination of positive and negative classes, was triggered by each observation’s hospital admission date. An observation was classified as positive if the corresponding patient had a recorded date of all-cause death within 1-year of the associated observation admission date. All other HF patient admission observations were classified as negative.

After the application of exclusion criteria, our final VHA V-CHAMPS HR validation set consisted of 75,086 HF patients. These patients sustained 380,441 hospital admissions that met our observation criteria. Of these, 85,956 (22.59%) were positive observations. Out of all the files used for model training, *lab_results_train* contributed to 258,527,315 EHR entries, *demographics_static_train* contributed to 133,252 EHR entries, *measurements_blood_pressure_train* contributed to 21,997,558 EHR entries, *measurements_train* contributed 125,247,162 EHR entries, *inpatient_admissions_train* contributed to 522,740 EHR entries, *medications_administered_train* contributed to 123,849,585 EHR entries, *conditions_train* contributed to 88,266,471 EHR entries, *procedures_train* contributed to 103,020,504 EHR entries, and *death_train* contributed to 97,284 EHR entries. A total of 721,661,871 EHR data entries were available for model training and development across all csv data files. The model predicted 1-year mortality with an AUC of 0.79.

### Feature Extraction and Deep Learning Network Design

As described in the previous report, the employed deep learning model (**Figure 1**) incorporates three long short-term memory (LSTM) layers to extract features from the time-series EHR data. Additionally, a dense layer processes non-time-series patient attributes such as age at admission, sex, race, and length of stay (LOS). Finally, an additional dense hidden layer further refines the features derived from the outputs of the three LSTM layers and the initial dense layer.

**Figure 1.**
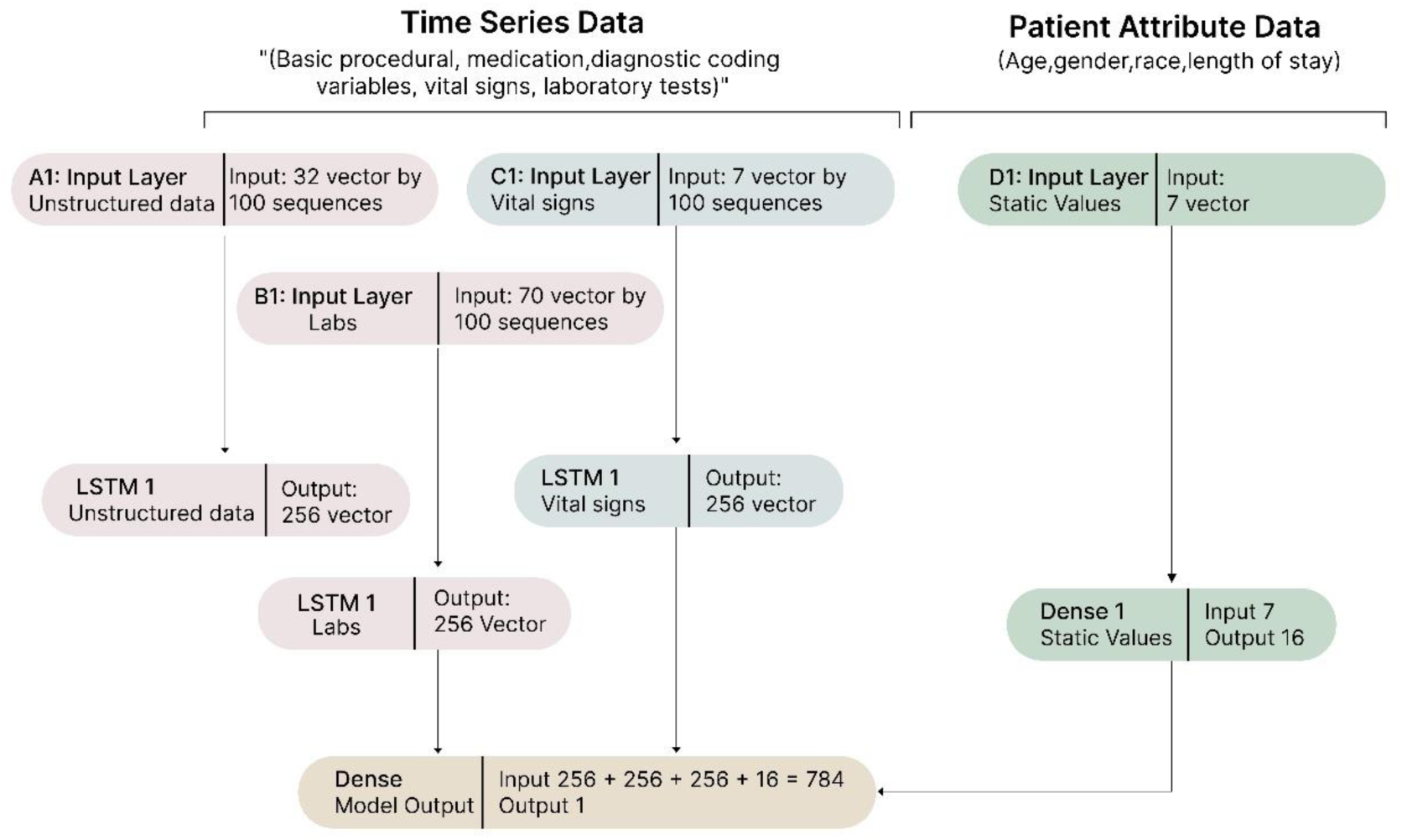
Ensemble Deep Learning Model Structure. Ensemble deep learning models were created using both time-series and densely connected networks, leveraging electronic health record (EHR) data. The model incorporates three long short-term memory (LSTM) layers to extract features from time-series EHR data. Additionally, a dense layer processes non-time-series patient attributes such as age at admission, sex, race, and length of stay (LOS). A final dense hidden layer further refines features based on the outputs from the three LSTM layers and the dense layer.

The model employs three LSTM layers to generate three intermediate feature vectors, each of length 256, from the time-series input data. Simultaneously, a dense input layer processes the non-time-series attributes (age, sex, race, and LOS) to produce an intermediate feature vector of length 16. These four vectors are then passed to a dense hidden layer, which consists of 16 neurons and an additional bias neuron. This final dense hidden layer integrates features from all previous layers and outputs a 16-length vector to the final output neuron, which performs the classification task. All dense hidden layers use the rectified linear unit (ReLU) activation function, while the final classification output neuron utilizes a sigmoid activation function.

Missing demographic data were managed as follows: For missing age, a default value of 61 (the mean age of the dataset) was used. Sex was encoded as 1 for male, −1 for female, and 0 for missing or unknown. Race was represented via one-hot encoding using four classes: White, Black, Asian, and Other. If race was unknown, all said variables were all set to zero. Records with missing demographic values were excluded from the training set to ensure they did not influence study results.

In order to incorporate the predictive power of patient-specific clinical information accumulated (in both inpatient and outpatient settings) before each observation hospital admission, the model used all available time-series EHR data leading up to each admission (up to a total of 100 visits), along with the EHR data actually collected during that admission’s hospital stay. Each of the model’s three LSTM layers processed sequences with a maximum length of 100 time-slices, representing the most recent 100 hospitalizations or outpatient clinic visits. If fewer than 100 time-slices were available, the missing values were filled with dataset mean values to ensure a complete data array.

Our encoding process produced the following tensor shapes: a 100 x 7 tensor for vital signs, a 100 x 71 tensor for laboratory test results, and a 100 x 32 tensor for diagnostic codes, medications and basic procedures. While outpatient EHR data is included in the 100 time slices of data accumulated before hospital admission, it cannot be used as an independent observation in the model without being linked to a hospital admission. The model is not designed to make predictions based solely on isolated outpatient EHR data.

The laboratory test model input (Table 1), encoded as a vector of 71 values, initially uses the dataset’s mean value for each test. As new laboratory test results are recorded, the tensor is updated with the latest values for *each of the corresponding dates*, retaining these results until new values are recorded. Consequently, abnormal lab results remain until replaced by different values in the patient’s EHR. Since laboratory tests have high cardinality, most patients only have a subset of the 71 identified test values.

**Table 1.** Impact of Specific Variables on Model Outcome Prediction.

| Variable Name in VHA V-CHAMPS Database | Variable Definition | AUC Without Variable | Importance |
| --- | --- | --- | --- |
| Age at admission | Age at admission (years) | 0.729498 | 1 |
| Systolic BP | Systolic blood pressure (mmHg) | 0.775004 | 1 |
| Alkaline phosphatase | Serum alkaline phosphatase (IU/L) | 0.781374 | 1 |
| Weight | Weight (lb) | 0.776498 | 0.993361 |
| Chloride | Serum chloride (mEq/L) | 0.783488 | 0.990329 |
| Carbon dioxide | Serum carbon dioxide (mmol/L) | 0.784893 | 0.983902 |
| Pulse | Heart rate (bpm) | 0.779907 | 0.978208 |
| Albumin | Serum albumin (g/L) | 0.786386 | 0.977075 |
| Calcium | Serum calcium (mg/dL) | 0.786543 | 0.976358 |
| International normalized ratio | International normalized ratio | 0.786689 | 0.97569 |
| Sodium | Serum sodium (mEq/L) | 0.787306 | 0.972866 |
| Glucose | Serum glucose (mg/dL) | 0.787479 | 0.972078 |
| Anion gap | Serum anion gap (mEq/L) | 0.787786 | 0.97067 |
| Prothrombin time | Prothrombin time (seconds) | 0.787904 | 0.97013 |
| Aspartate aminotransferase | Serum aspartate aminotransferase (IU/L) | 0.787948 | 0.969928 |
| Potassium | Serum potassium (mEq/L)<br>*\*Laboratory test* | 0.787954 | 0.969902 |
| Urine specific gravity | Urine specific gravity | 0.787974 | 0.969811 |
| Brain natriuretic peptide | Serum brain natriuretic peptide (pg/mL) | 0.787987 | 0.969754 |
| Diastolic BP | Diastolic blood pressure (mmHg) | 0.781814 | 0.969734 |
| Fasting glucose | Fasting serum glucose (mg/dL) | 0.787993 | 0.969722 |
| Partial thromboplastin time | Partial thromboplastin time (seconds) | 0.788014 | 0.969628 |
| Thyroid stimulating hormone | Thyroid stimulating hormone (μU/mL) | 0.78807 | 0.969371 |
| Partial pressure of carbon dioxide | Partial pressure of carbon dioxide (mmHg) | 0.788076 | 0.969343 |
| Methemoglobin | Methemoglobin (% of hemoglobin) | 0.788164 | 0.968943 |
| Fibrinogen | Fibrinogen (mg/dL) | 0.788173 | 0.968903 |
| Bicarbonate | Serum bicarbonate (mmol/L) | 0.788207 | 0.968746 |
| Creatinine | Serum creatinine (mg/dL) | 0.788212 | 0.968725 |
| Lactate dehydrogenase | Serum lactate dehydrogenase (IU/L) | 0.788229 | 0.968646 |
| Lipase | Serum lipase (IU/L) | 0.788234 | 0.968624 |
| Direct bilirubin | Direct bilirubin (mg/dL) | 0.788243 | 0.968579 |
| D-dimer | D-dimer | 0.78825 | 0.968549 |
| C-reactive-protein | C-reactive-protein (mg/L) | 0.788262 | 0.968495 |
| Cholesterol | Serum cholesterol (mg/dL) | 0.788267 | 0.96847 |
| Lactic acid | Serum lactic acid (mmol/L) | 0.788288 | 0.968375 |
| Uric acid | Serum uric acid (mg/dL) | 0.788301 | 0.968314 |
| Alanine aminotransferase | Alanine aminotransferase (U/L) | 0.78831 | 0.968273 |
| Magnesium | Serum magnesium (mg/dL) | 0.788313 | 0.968259 |
| Ammonia | Serum ammonia (µg/dL) | 0.788318 | 0.968238 |
| Amylase | Serum amylase (IU/L) | 0.788321 | 0.968226 |
| Bilirubin | Total bilirubin (mg/dL) | 0.788329 | 0.968189 |
| Plasma protein | Plasma protein (g/dL) | 0.788334 | 0.968163 |
| Neutrophils | Neutrophils (cells/µL) | 0.788337 | 0.968152 |
| Hemoglobin A1c | Hemoglobin A1c (%) | 0.78834 | 0.968136 |
| Monocytes | Monocytes (cells/µL) | 0.788342 | 0.968126 |
| Oxygen gradient | Alveolar-arterial oxygen gradient (mmHg) | 0.788359 | 0.96805 |
| Oxygen saturation | Arterial oxygen saturation (%) | 0.788364 | 0.968029 |
| Partial pressure of oxygen | Arterial partial pressure of oxygen (mmHg) | 0.788366 | 0.968019 |
| Troponin I | Troponin I (ng/mL) | 0.788382 | 0.967947 |
| Creatine kinase | Creatine kinase (IU/L) | 0.788385 | 0.967933 |
| Platelets | Platelets (cells/µL) | 0.788392 | 0.967899 |
| Eosinophils | Eosinophils (cells/µL) | 0.788395 | 0.967887 |
| pH | pH | 0.788402 | 0.967853 |
| Pulmonary artery oxygen saturation | Pulmonary artery oxygen saturation (%) | 0.788409 | 0.967821 |
| Plasma potassium | Plasma potassium (mEq/L)<br>*\*Point-of-care test* | 0.788409 | 0.96782 |
| Calculated oxygen saturation | Calculated arterial oxygen saturation (%) | 0.788409 | 0.96782 |
| Pulmonary artery oxyhemoglobin | Pulmonary artery oxyhemoglobin (%) | 0.788409 | 0.96782 |
| Indirect coombs | Indirect coombs (binary) | 0.788409 | 0.96782 |
| Hemoglobin total | Hemoglobin total (g/dL)<br>*\*Point-of-care test* | 0.788409 | 0.96782 |
| Cocaine | Urine cocaine metabolites (binary) | 0.78842 | 0.967772 |
| Basophils | Basophils (cells/µL) | 0.788423 | 0.967757 |
| Hematocrit | Hematocrit (%) | 0.788425 | 0.96775 |
| Plasma phosphorus | Plasma phosphorus (mg/dL) | 0.788451 | 0.96763 |
| FiO2 | Fraction of inspired oxygen (%) | 0.788452 | 0.967626 |
| Carboxyhemoglobin | Carboxyhemoglobin (%) | 0.788464 | 0.967568 |
| Lymphocytes | Lymphocytes (cells/μL) | 0.788473 | 0.967531 |
| Triglycerides | Serum triglycerides (mg/dL) | 0.788473 | 0.967528 |
| High density lipoprotein | Serum high density lipoprotein (mg/dL) | 0.788487 | 0.967467 |
| Base excess | Base excess (nmol/L) | 0.788514 | 0.967343 |
| Ketones | Plasma ketones (mmol/L) | 0.78854 | 0.967223 |
| Hemoglobin | Hemoglobin (g/dL)<br>*\*Laboratory test* | 0.788573 | 0.967069 |
| Mixed venous oxygen saturation | Mixed venous oxygen saturation (%) | 0.788614 | 0.966885 |
| Benzodiazepines | Urine benzodiazepines (binary) | 0.788616 | 0.966874 |
| Blood urea nitrogen | Blood urea nitrogen (mg/dL) | 0.788635 | 0.966787 |
| Cannabinoids | Urine cannabinoids (binary) | 0.78866 | 0.966672 |
| Mixed venous oxygen | Mixed venous oxygen (partial pressure; mmHg) | 0.788928 | 0.965448 |
| O2 saturation | Oxygen saturation (via pulse oximetry; %) | 0.787489 | 0.944509 |
| Height | Height (in) | 0.788283 | 0.940982 |
| O2 flow rate | Oxygen flow rate (L/min) | 0.788409 | 0.94042 |
| - | Unstructured row count | 0.781932 | 0.806163 |
| Race | Race (categorical) | 0.784651 | 0.796109 |
| Length of stay hours | Length of stay (hours) | 0.785356 | 0.793505 |
| - | Vital sign row count | 0.787143 | 0.786896 |
| Gender | Sex (categorical) | 0.787477 | 0.785663 |
| - | Laboratory test row count | 0.787534 | 0.785452 |

Abbreviations: AUC, area under the curve (for a receiver operating characteristic curve); bpm, beats per minute; dL, deciliter; g, grams; in, inches; IU, international units; L, liter; lb, pounds; mEq, milliequivalents; mg, milligrams; mL, milliliter; min, minute; mmHg, millimeters of mercury; mmol, millimole; ng, nanograms; nmol, nanomole; pg, picogram; $\mu$ L, microliter; $\mu$ U, microunits.
\*Where laboratory test or point-of-care test is specified, the variable in question was one of two variables in the VHA V-CHAMPS database that both represented the results of two different tests for the same substance (e.g. potassium), where one test was sent to and performed in a laboratory, and the other was a point-of-care/bedside test.

Similarly, vital signs (Table 1) are encoded as a vector of 7 values. Missing vital sign data are filled with dataset mean values for each specific vital sign. Each column in the sequence represents either the most recent vital sign value or the dataset means if no recent value is available. Notably, the time slices for vital signs reflect the mean values for each of the past 100 EHR clinical encounters.

All basic procedural, medication, and diagnostic coding variables were mapped to 100-length sequences within a 32-dimensional vector space (generated using Python’s Gensim Word2Vec) before being input into the LSTM layer [27]. These variables are high-cardinality and sparse. Since the data was encoded into 32-dimensional vectors that are updated over time, there was no need to explicitly handle missing data. Only present values are encoded into the time series; values that are not present are simply excluded and not represented in the sequence.

The primary metric for evaluating model performance is the area under the receiver operating characteristic (ROC) curve (AUC). Additional performance metrics include log loss, precision-recall (PR) AUC, and Brier score (Table 2). Feature importance was assessed using the permutation feature importance method [18], which involves evaluating the impact of permuting specific input features on the AUC of a trained neural network.

**Table 2.** Measures of Model Performance.

| Measure | Result |
| --- | --- |
| ROC AUC<br>(Area under the receiver operating characteristic curve) | 0.79 |
| Log Loss | 0.49 |
| PR AUC<br>(Area under the precision recall curve) | 0.53 |
| Brier Score | 0.15 |

When training deep learning models on heterogeneous EHR data, variations in data structure and clinical and financial decision-making between institutions mean that model weights learned from one hospital’s data cannot be assumed to generalize to another. Consequently, model validation here reflects validation of the underlying architecture, as opposed to the architecture and the associated weights. This involves retraining the model weights using data from the new institution to assess whether the architecture itself has predictive power in the new context, rather than relying on weights derived from a different, potentially incompatible data source.

## Results

Table 3 presents the demographic characteristics of all study observations, Supplementary Table 1 compares them to the demographics of the original model development dataset. Among the 380,441 observations, 85,956 (22.59%) were labeled as positive and 294,485 (77.41%) as negative, based on our classification criteria. The average age of patients at the time of observation was 69 years, with the majority being white and male. Table 4 provides descriptive statistics for positive and negative observations, including mean values for LOS hours, counts of vital sign data, laboratory values, and unstructured data, and Supplementary Table 2 compares the same to those for the original model development dataset.

**Table 3.** Observation Population Demographics.

| Model Observations | Negative | Positive | Total |
| --- | --- | --- | --- |
| Number of observations, *n* (%) | 294,485 (77) | 85,956 (23) | 380,441 |
| Age at observation (years), *mean* (*SD*) | 68 (11) | 75 (11) | 70 (12) |
| Sex of observation patient, *n* (%) |  |  |  |
| Male | 284,410 (96.4) | 84,225 (97.7) | 368,635 (96.9) |
| Female | 10,075 (3.4) | 1,731 (2.0) | 11,806 (3.1) |
| Race of observation patient, *n* (%) |  |  |  |
| White | 63,793 (21.7) | 15,182 (17.7) | 78,975 (20.8) |
| Black | 207,533 (70.5) | 57,461 (66.8) | 264,994 (69.7) |
| Asian | 836 (0.3) | 324 (0.4) | 1,160 (0.3) |
| Other | 22,323 (7.6) | 12,989(15.1) | 35,312 (9.3) |

**Table 4.**
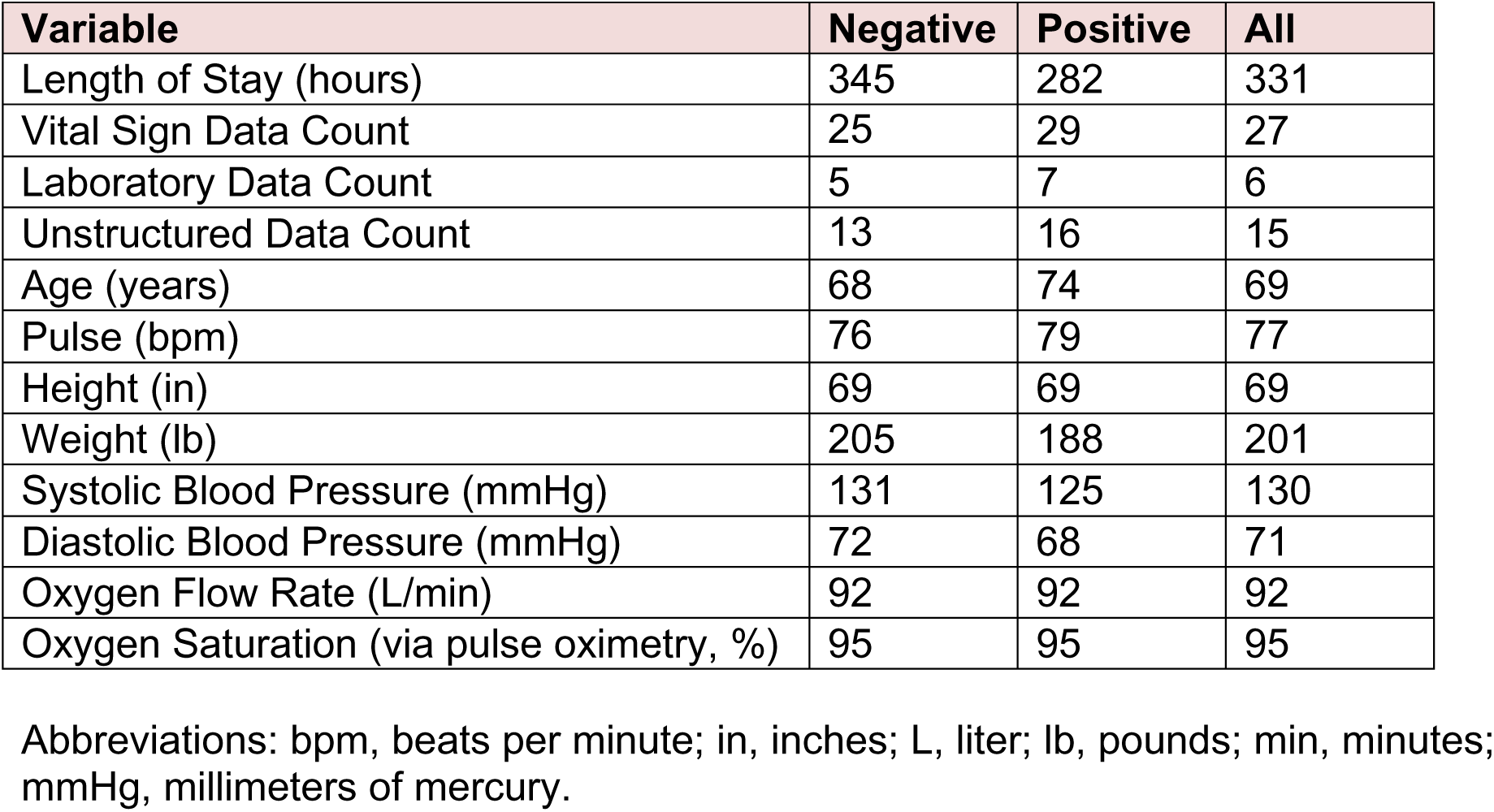
Variable Means by Positive and Negative Observation Groups.

Table 1 highlights the univariate importance of various features, including vital signs, laboratory tests, and static variables. Among these, serum albumin, age at admission, systolic BP, serum alkaline phosphatase, serum carbon dioxide, international normalized ratio, and arterial partial pressure of CO_2_ emerged as the most significant features.

Multivariate analysis of feature importance can reveal intricate relationships within EHR data, potentially predicting responses to medical treatments and generating mechanistic hypotheses. Figure 2 presents a heat map illustrating the interaction between vital sign and laboratory test variables. The x-axis and y-axis represent vital sign and laboratory test features, respectively, ordered by their univariate importance. The heat map highlights the significant impact of cocaine use and weight on model predictions.

**Figure 2.**
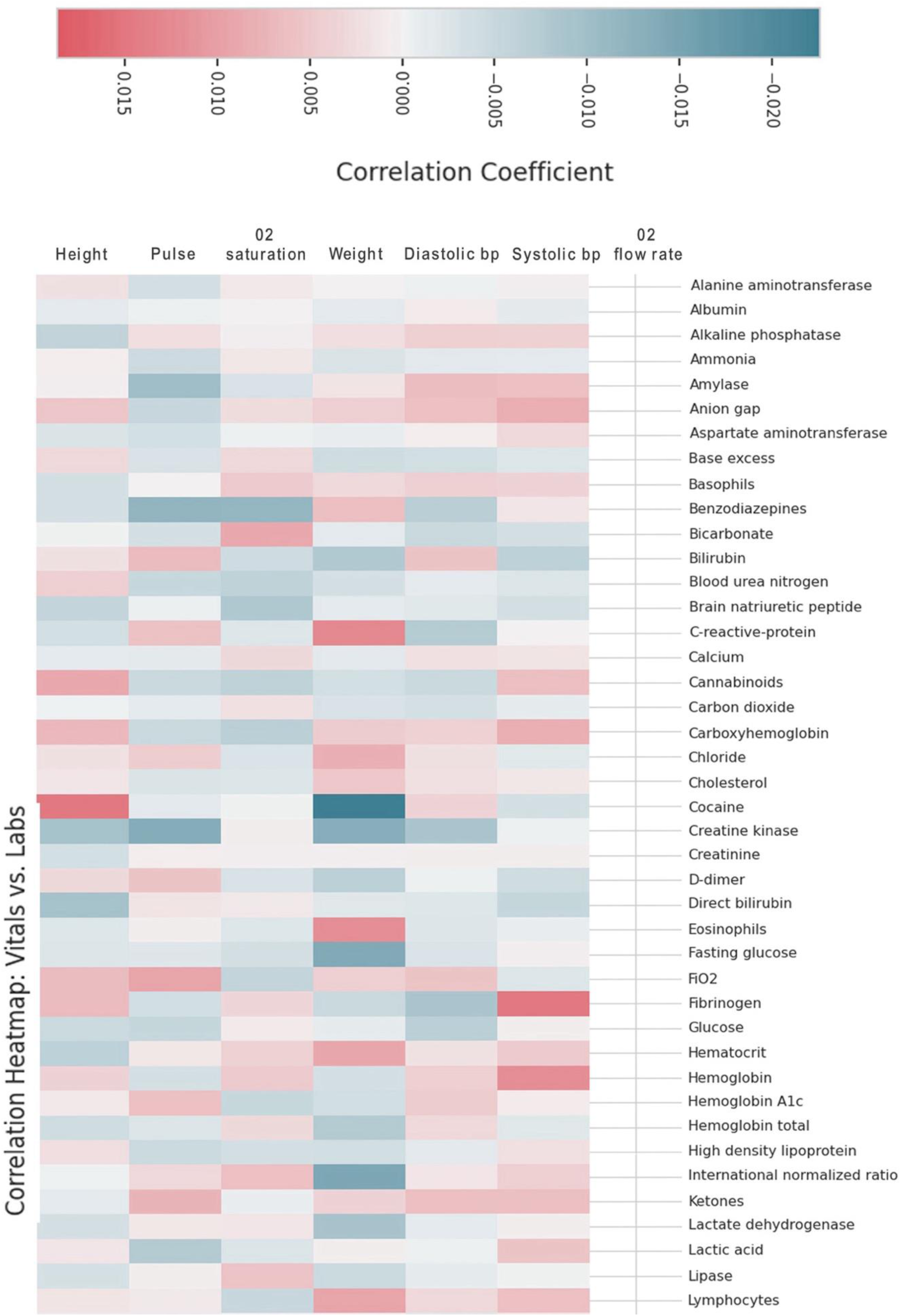
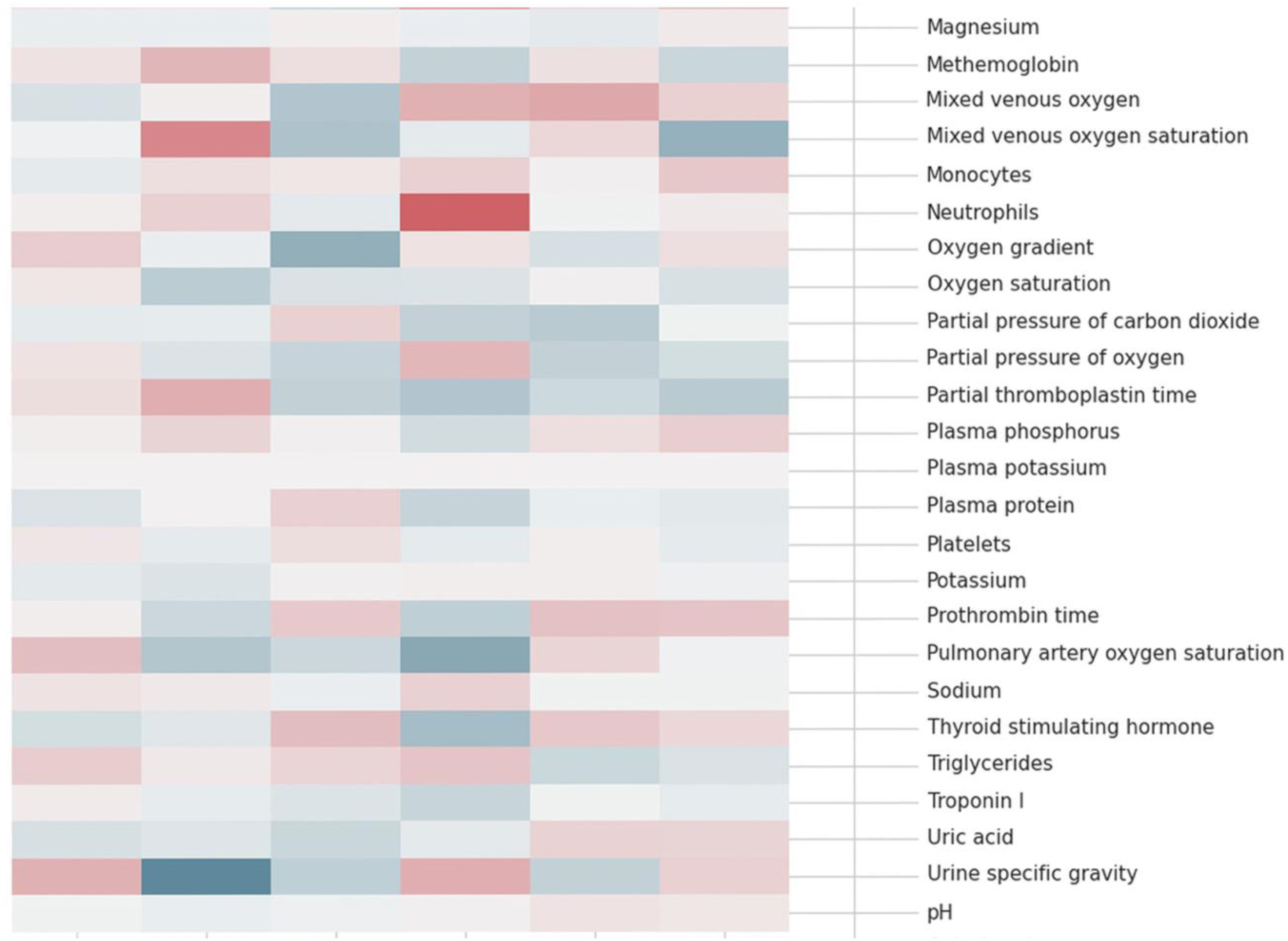
Heat Map of Vital Sign Versus Laboratory Test Values. This heat map illustrates the interaction between vital sign and laboratory test variables, with vital signs on the x-axis and laboratory values on the y-axis. Variables are arranged according to their univariate importance. Cocaine and Weight have the most significant impact on model predictions. Abbreviations: AG, anion gap; Alk Phos, alkaline phosphatase; ALT, alanine aminotransferase; AST, aspartate aminotransferase; Base Ex, base excess; Benzos, benzodiazepines; Bicarb, bicarbonate; BNP, brain natriuretic peptide; BUN, blood urea nitrogen; Chol, cholesterol; CK, creatine kinase; CO2, carbon dioxide; COHb, carboxyhemoglobin; CRP, c-reactive protein; Diastolic, diastolic blood pressure; Dir Bili, direct bilirubin; fiO2, fraction of inspired oxygen; Gluc Fast, fasting glucose; HbA1c, hemoglobin A1c; Hct, hematocrit; Hgb, hemoglobin; Hgb total, hemoglobin total; HDL, high-density lipoprotein; Ind Coombs, indirect Coombs; INR, international normalized ratio; LDH, lactate dehydrogenase; Mg, magnesium; MetHb, methemoglobin; Na, sodium; O2 Flow, oxygen flow; O2 Sat, oxygen saturation; O2 Sat Cal, calculated oxygen saturation; O2 Sat ven, mixed venous oxygen saturation; OxygenPA, pulmonary artery oxygen saturation; Oxy grad, oxygen gradient; OxyhemePA, pulmonary artery oxyhemoglobin; pCO2, partial pressure of carbon dioxide; pO2a, arterial oxygen partial pressure; pO2v, mixed venous oxygen partial pressure; Plasma K, plasma potassium; PT, prothrombin time; PTT, partial thromboplastin time; Spec Grav, urine specific gravity; Systolic Bp, systolic blood pressure; TSH, thyroid stimulating hormone; Trigly, triglycerides.

### Model Performance Evaluation

Figure 3 shows the validation model performance by ROC curve. The AUC for the deep learning model test set was 0.79 with 95% confidence interval of (0.78-0.79). The AUC for our training set was 0.79. We utilized a validation set to determine when to stop training. This validation set had an AUC of 0.79.

**Figure 3.**
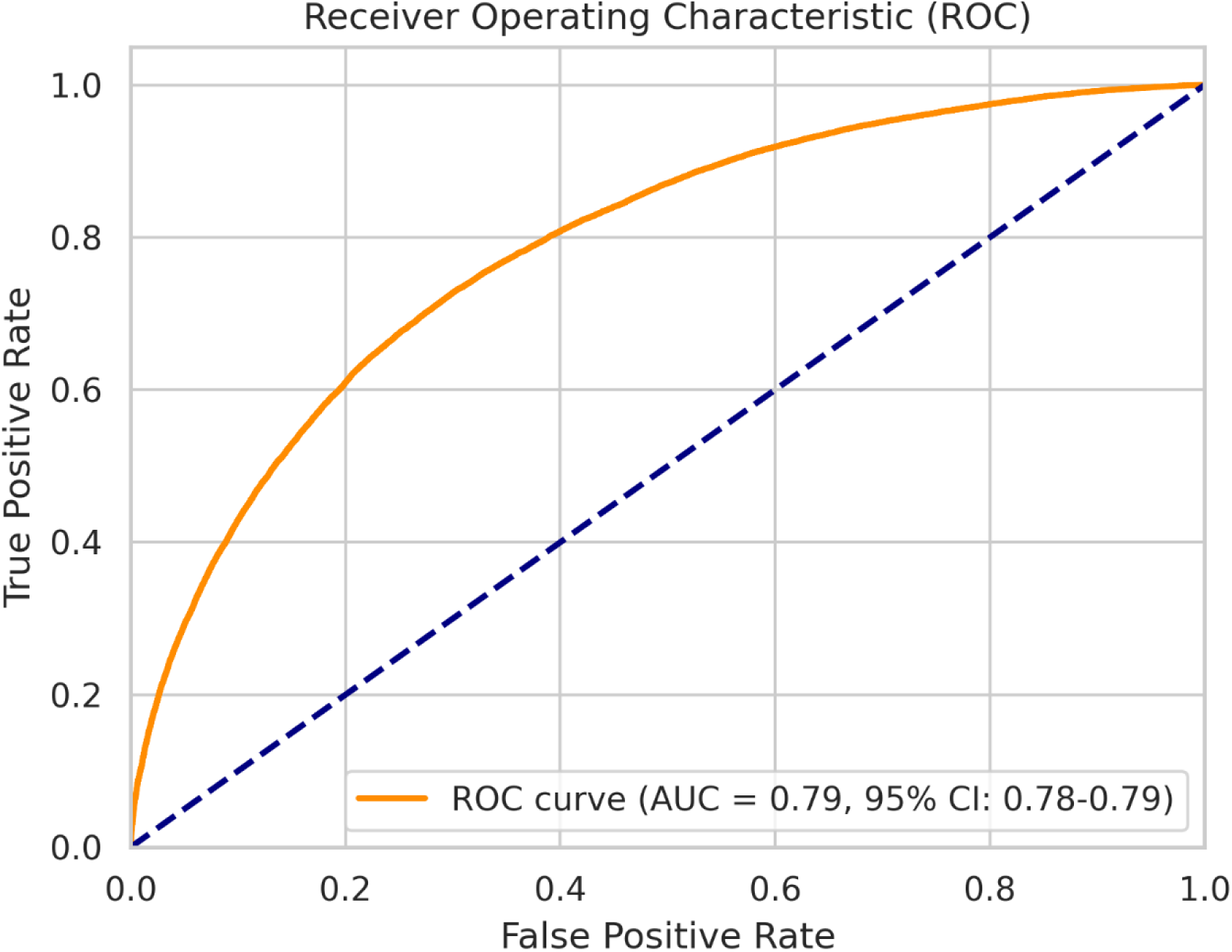
Receiver Operating Characteristic (ROC) Curve for the Deep Learning Model’s Prediction Performance. The ROC curve’s area under the curve (ROC AUC or C-statistic) of 0.79 with a confidence interval of 0.78 to 0.79 supports deep learning model clinical relevancy.

The accuracy and loss values for the training and test sets were 75.87 and 0.54, respectively. The validation accuracy was 78.84 and the validation loss was 0.49. The AUC was 0.79. The precision, recall, and other metrics are shown in Table 2. The precision recall (PR) area under the curve (PR AUC) was 0.53 with a CI of 0.53 to 0.55. The PR AUC 0.53 referenced in Table 2 is shown in Figure 4.

**Figure 4.**
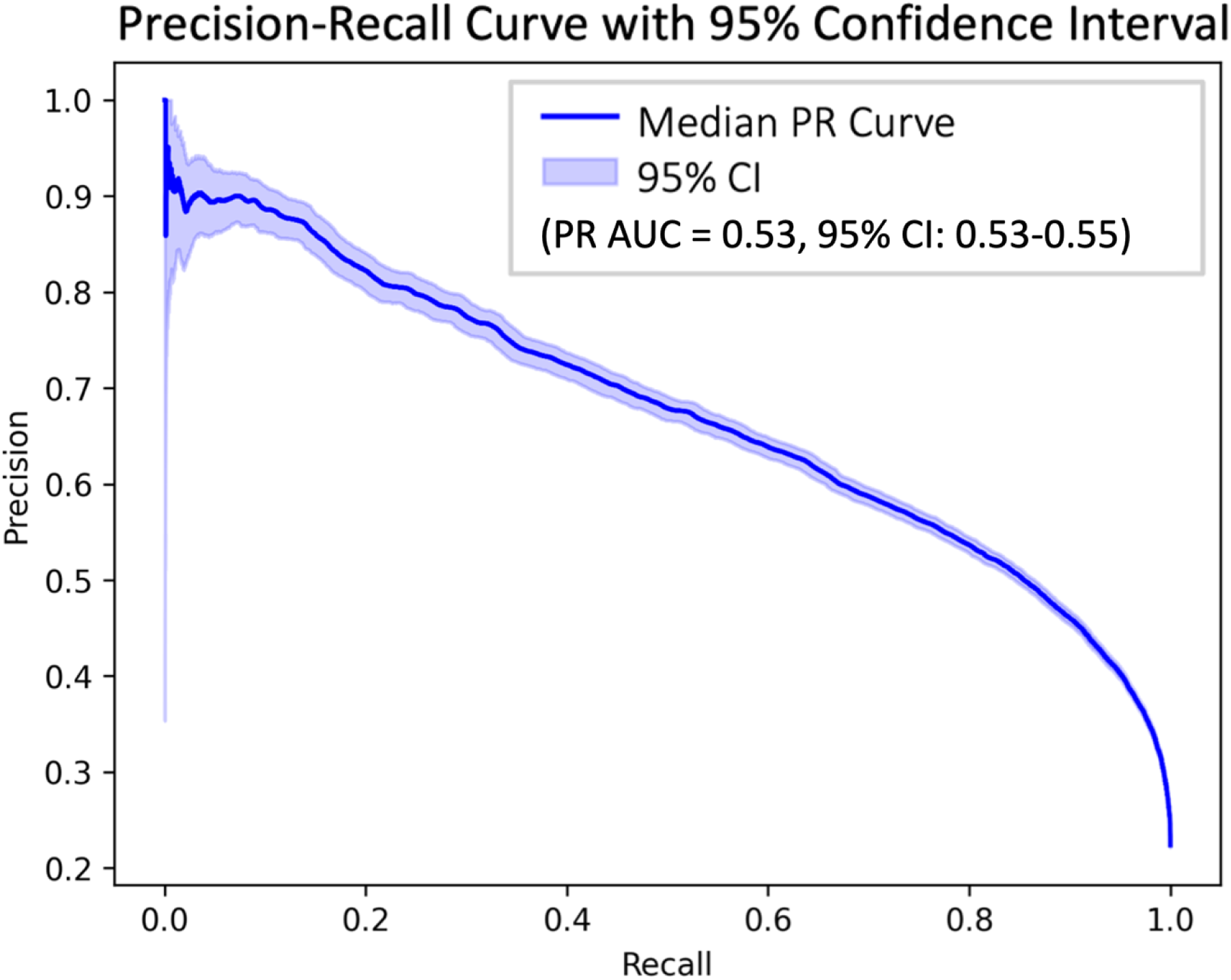
Precision Recall (PR) Curve performance metric. The PR area under the curve (PR AUC) of 0.53 with a confidence interval of 0.53 to 0.55 further supports the clinical relevancy of our predictive deep learning model.

**Figure 5.**
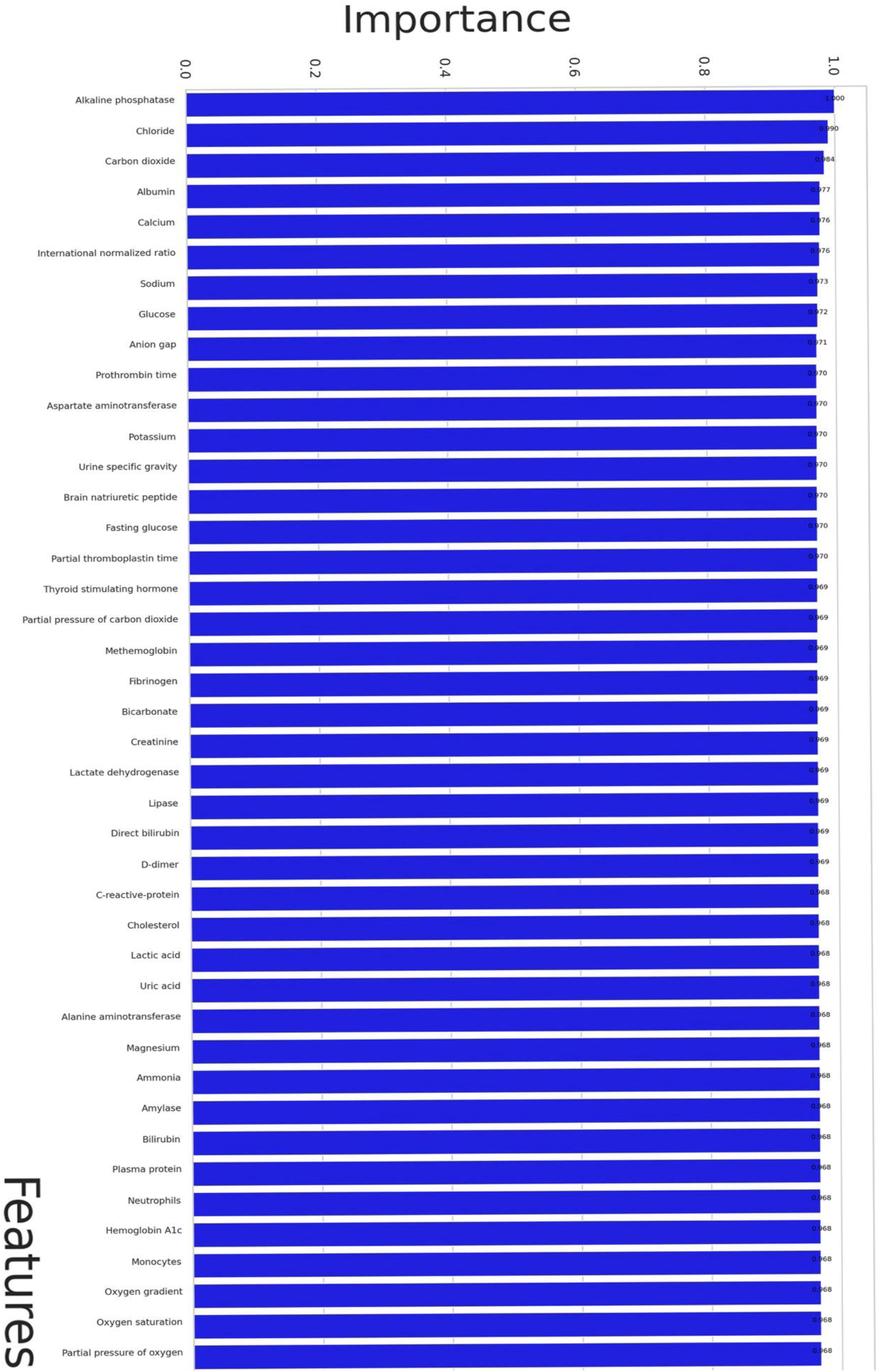
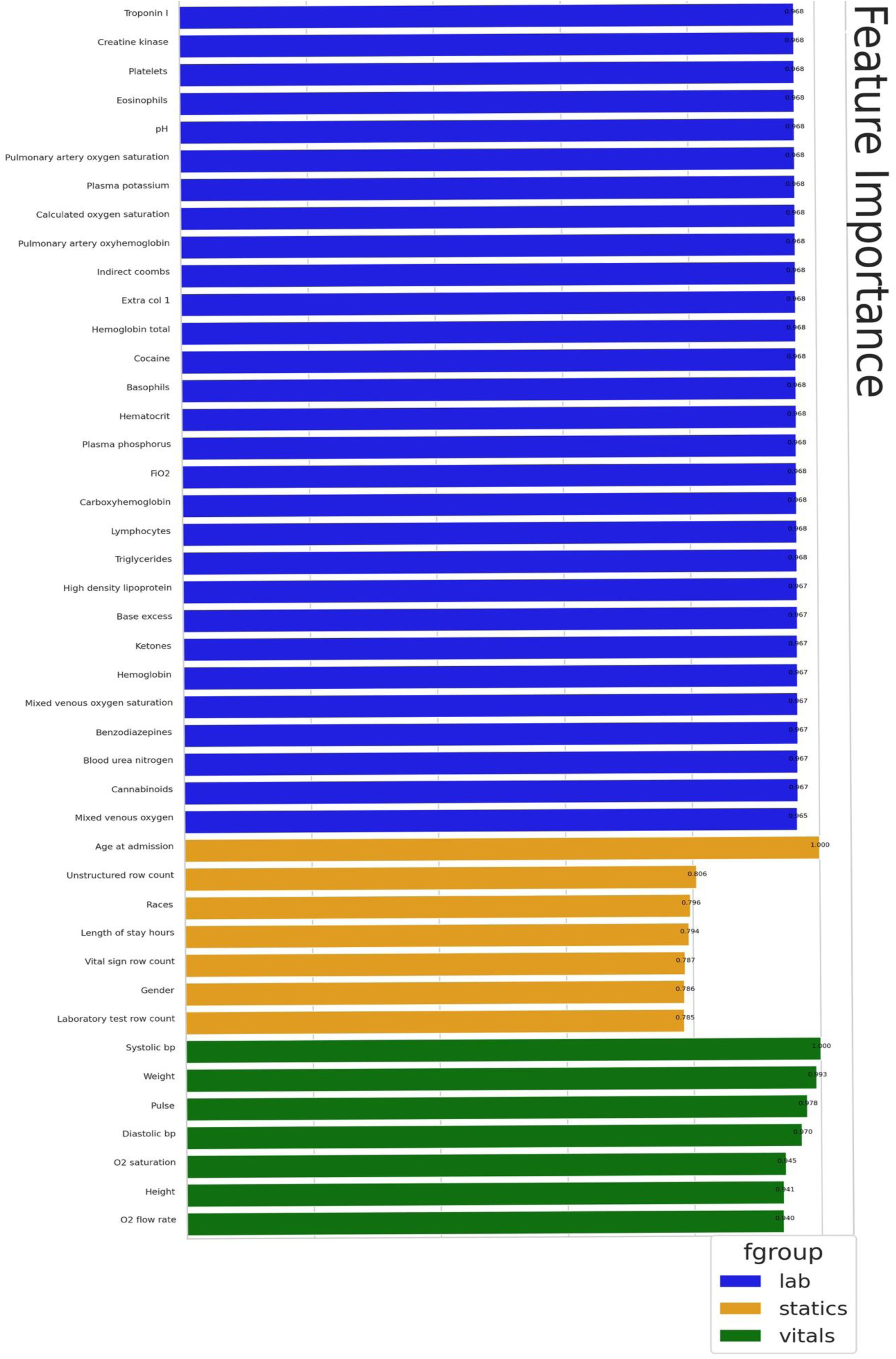
Feature Importance. Vital sign variables, laboratory variables, and static variables impacting our predictive deep learning model. Age at admission, weight, systolic blood pressure, serum alkaline phosphatase, serum chloride, and serum carbon dioxide emerged as the most significant features.

## Discussion

Outcome prediction continues to be a foundational component of HF patient care, directly impacting patient management decisions. Most of the therapeutic choices involving the use of high-risk invasive HF interventions are persuasively influenced by near-term clinical risk prediction. To this end, there is considerable evidence supporting the predictive potential of deep learning models when challenging clinical scenarios such as this are aligned with appropriate problem/data-specific algorithms [29].

In this regard, the exceptional ability of deep learning models to parse innumerable complex predictive relationships between many widely differing—and often unpredictable—clinical variable combinations offers unique potential to assist clinicians in difficult HF management decisions. The goal of deep learning application in these specific scenarios is to improve the accuracy of clinician identification of those HF patients whose predicted near-term risk defines them as having the most to gain from invasive interventions.

Our approach to model validation here focusses on validating the underlying model architecture (that is, the structure and attributes of the underlying LSTM layers and nodes) as opposed to assuming transferrablity of model architecture and weights from one clinical setting to another. In a perfect world, all EHRs would be 1:1 comparable across different insitutions or datasets, meaning model archiectures and weights trained on one source could be applied “out of the box” to another. In practice, such an assumption cannot be made. Differences in procedural reporting, data structure, and clinical practices may mean that numerically similar numbers or classifications have different clinical meanings. Because deep learning models enable a highly non-linear transformation of input to output, even small changes in the underlying assumptions associated with input data may have far-reaching (and potentially misleading) consequences for output. To mitigate this, our approach here focused on validating the underlying model architecture rather than assuming transferrablity of both the architecture and the trained weights. Practically speaking, this means if our model were to be applied in a clinical context, one would need to train new weights based on extant data from that context prior to model deployment, building a bespoke set of weights that work alongside a generalized model architecture. As such, we consider ease and computational resources needed for model training (as well as prediction) to be of major practical consideration.

Our model makes use of a series of long short-term memory (LSTM) layers. Recent progress in deep learning has turned to favor transformer-based archiectures for language processesing, especially in the context of large language models (LLMs)[19]. Compared to LSTM-based models, transformer-based LLMs leverage self-attention to learn longer-range correlations in context which are often lost in ‘simpler’ archictures. While this can be powerful for more complex patterns of data, these models come with a substantial computational cost for both training and prediction (inference)[20]. While the transition to transformer-based archiectures away from LSTM-based deep learning models may seem like a logical next step, careful consideration of the tradeoffs between accuracy and performance given the training data will be important, especially when EHR data may often lack the types of long-range complexity for which transformers are particularly well-suited. Moreover, as discussed above, if model deployment requires training on institution-specific datasets first, training large transformer-based models may be challenging as datasets scale, further highlighting the advantages of LSTM-based archiectures.

Although many EHR-based outcome predictive deep learning models have been developed in large HF patient datasets, few of these model architectures have been subjected to subsequent rigorous external validation in similarly large HF patient datasets not used in the original model development. While our original EHR HF model was based upon 79,850 observation admissions from 52,265 HF patients and achieved an AUC of 0.91 in predicting 1-year mortality or severe decompensation, generalization to other HF datasets cannot be assumed. Validation of these results in external HF populations not used in model training is essential before clinical application.

This investigation reports follow-up external validation of our previously reported single-center, EHR-based deep learning model in a similarly large HF patient EHR *validation* test set. This external validation test set is provided by the VHA V-CHAMPS HF challenge and includes 380,441 admission observations contributing 721,661,871 EHR variables from 75,086 HF patients. This particular external validation test set also critically examines model generalization to the extremes of the United States HF population by its unique inclusion of patients from every state in the country. The validation test set results reported in this current investigation again support our model’s clinical relevance in 1-year HF outcome prediction with a model AUC of 0.79.

Although some mixed results have been reported [21–24], most previously reported investigations support the hypothesis that strong outcome predictive relationships exist in EHR data acquired during routine HF patient management. Moreover, there is an ever-growing appreciation that this predictive power is exploitable by deep learning models to predict real-world non-response to HF medical therapy [25–27]. These EHR-based deep learning models may therefore have considerable potential to assist clinicians in the more accurate direction of highly invasive, life-disruptive, and yet potentially life-saving therapy in end-stage HF patients.

The confirmed accuracy and generalizability of our model may hinge upon its use of machine learning principles and archiectures that are adept at capturing the predictive power inherent in the associated temporal patterns of time-sensitive EHR data. This approach may allow for the detection of even subtle trends in HF disease progression or remission that are reflected in the temporal evolution of EHR-based metrics. The use of temporal information also aligns intuitively with longstanding HF clinical management paradigms. Specifically, our model utilizes machine learning architectures developed to capture time-series data [22,26,30] to analyze all variables except for non-time-dependent, fixed variables like demographics. Rather than using snapshots that report on a patient’s status at a specific moment, the ability to examine trends across multiple temporal observables may be the key component that enables a high degree of model accuracy. This investigation also supports the notion that EHR-based patient attribute descriptions resident in large HF datasets remain outcome predictive whether the patient population is from our large academic medical center or from the equally large VHA V-CHAMPS HR dataset drawn from patients across the US.

Applications that readily interact with existing EHR systems can enable the further realization of the full clinical potential of deep learning EHR-based outcome. Most EHR systems, including EPIC and Cerner, support application programming interfaces (APIs) that utilize the HL7 *Fast Healthcare Interoperability Resources* (FHIR) format. These APIs enable the automatic integration of patient-specific EHR data into EHR-embedded deep learning models. As these models progress, they have potential to provide clinicians with immediate patient-specific predictions of heart failure outcomes, thereby enhancing real-time patient management decisions.

### Limitations

This investigation has several potential limitations. These include its use of retrospective EHR data from many VHA institutions, with the well-recognized difficulty in assuring uniformity in data quality control across a large number of HF data acquisition sites scattered across a large geographical area. Further, all deep learning models using retrospective EHR data are susceptible to errors in temporal registration of time-based data, such that the model may inadvertently include late, post-outcome indicators of subsequent HF classification endpoints that would not have been available to clinicians at the time of clinical management decisions. These errors can obviously bias the model and falsely enhance model accuracy. The preparation, deployment, and extensive use of the V-CHAMPS competition data[17] reasonably assures that the VHA data has been meticulously reviewed by many investigative groups for evidence of systematic leakage of endpoint information in both structured and unstructured EHR data.

Accurately determining medical cause of death within large investigative subpopulations always poses significant challenges. Our target HF patient population consists of individuals with complex medical profiles and multiple disease processes that may contribute to mortality risk. These are often difficult to disentangle with any degree of clinical precision. Defining our positive class using “all-cause” mortality, while pragmatic, probably includes deaths not directly attributable to HF. While imperfect, using all-cause mortality remains the most viable, clinically relevant option for this investigation.

## Conclusion

Accurate identification of HF patients who are within 1-year of death or severe decompensation would improve precision in application of advanced medical and surgical HF interventions. Our previously published EHR-based deep learning model was developed to predict 1-year death or referral for end-stage HF surgical intervention in a large cohort of patients and clearly demonstrated accuracy that supports clinical relevance. This clinical relevance in 1-year mortality prediction was confirmed in the present external validation study in the similarly large VHA V-CHAMPS HF dataset. Although many such HF outcome predictive models have been developed, few have been subjected to subsequent rigorous *external validation* in large HF patient datasets not used in model training.

These HF outcome predictive models have considerable potential for even further improvement in accuracy when the complex information residing in the vast quantity of unstructured EHR data is included. Further, the myriad of complex HF physiological, functional [25,30], and high-definition regional contractile indices [32,33], all of which hold the potential to instill even more clinical relevance, can be expected to enhance future model development. As these models continue to improve, automated EHR-embedded deep learning systems may allow direct, real-time assistance of clinicians in difficult HF patient management decisions.

## Data Availability

VHA Synthetic Data Lake (V-CHAMPS)

https://www.data.va.gov/stories/s/How-to-Access-Synthetic-Data-in-the-VA/rssm-v4rt/

## List of Abbreviations

API: application programming interfaces
AUC: area under the curve
BNP: brain natriuretic peptide
BP: blood pressure
ECMO: extracorporeal membrane oxygenation
EF: ejection fraction
EHR: electronic health record
FHIR: fast healthcare interoperability resources
HF: heart failure
LOS: length of stay
LSTM: long short-term memory
PR: precision recall
ROC: receiver operating characteristic
VAD: ventricular assist device

---

**Central Illustration.**
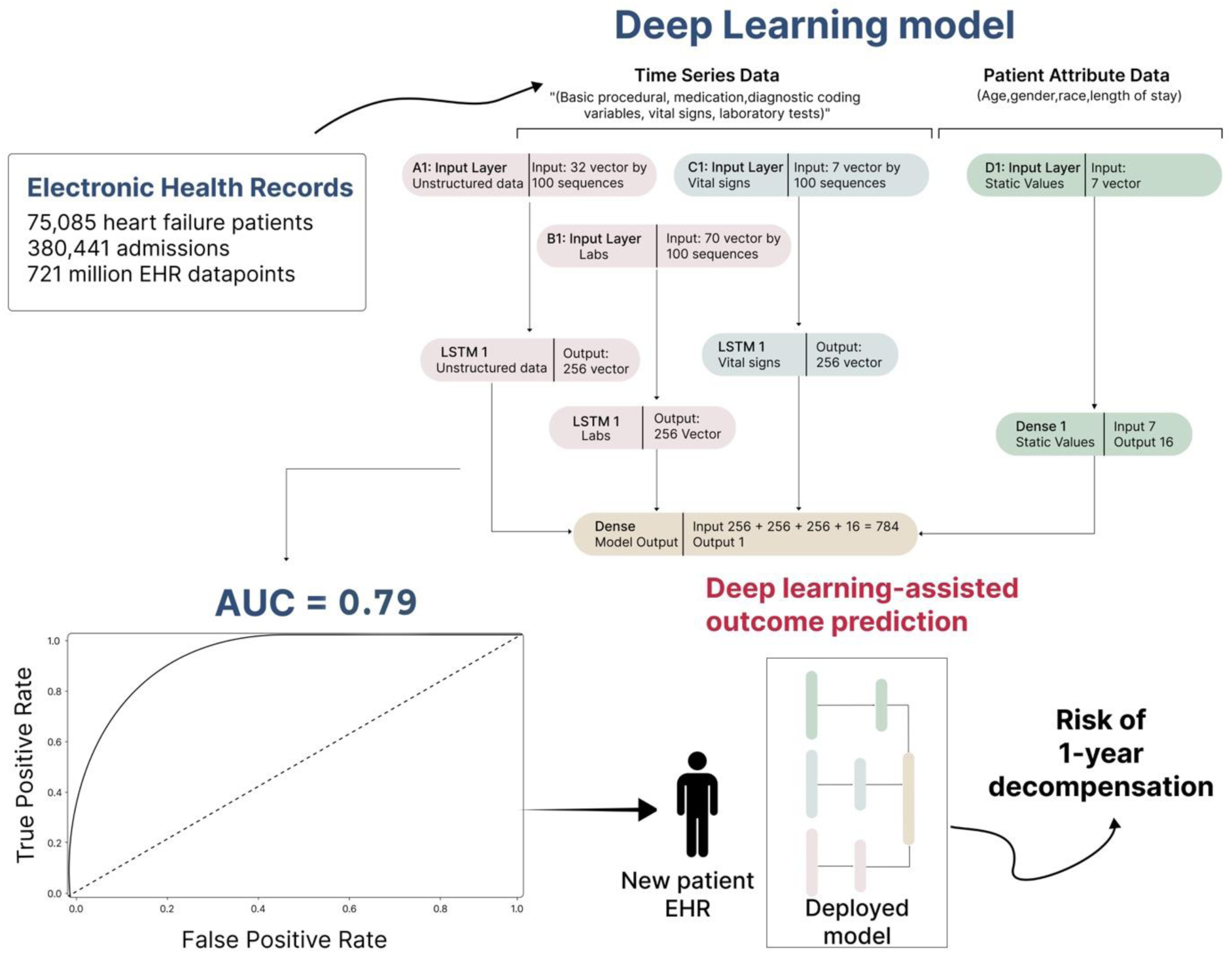
External validation of our previous electronic health record (EHR)-based deep learning model prediction of near-term heart failure mortality. External validation of a previously published (*JACC HF*) EHR-based deep learning model in a similarly large HF patient validation test set confirmed clinical relevancy in predicting 1-year mortality in heart failure patients. The model’s receiver operating characteristic curve (ROC) in this new validation test set had an area under the curve (AUC) of 0.79, further supporting clinical relevance. Abbreviation: LSTM, long short-term memory.

## Supplementary

**Supplementary Table 1.** Population Demographics of the Original Model Development and Validation Datasets.

| Model Observations | Validation Dataset |  |  | Original Model Development Dataset |  |  |
| --- | --- | --- | --- | --- | --- | --- |
|  | Negative | Positive | Total | Negative | Positive | Total |
| Number of observations, *n* (%) | 294,485 (77) | 85,956 (23) | 380,441 | 64,017 (80.2) | 15,833 (19.8) | 79,850 |
| Age at observation (years), *mean* (*SD*) | 68 (11) | 75 (11) | 70 (12) | 61 (15) | 65 (14) | 62 (15) |
| Sex of observation patient, *n* (%) |  |  |  |  |  |  |
| Male | 284,410 (96.4) | 84,225 (97.7) | 368,635 (96.9) | 33,981 (53.1) | 9,168 (57.9) | 43,149 (54.0) |
| Female | 10,075 (3.4) | 1,731 (2.0) | 11,806 (3.1) | 30,034 (46.9) | 6,661 (42.1) | 36,695 (46.0) |
| Race of observation patient, *n* (%) |  |  |  |  |  |  |
| White | 63,793 (21.7) | 15,182 (17.7) | 78,975 (20.8) | 25,233 (39.4) | 7,139 (45.1) | 32,372 (40.5) |
| Black | 207,533 (70.5) | 57,461 (66.8) | 264,994 (69.7) | 25,936 (56.1) | 6,752 (42.6) | 42,688 (53.5) |
| Asian | 836 (0.3) | 324 (0.4) | 1,160 (0.3) | 313 (0.4) | 78 (0.5) | 391 (0.5) |
| Other | 22,323 (7.6) | 12,989 (15.1) | 35,312 (9.3) | 2,535 (4.1) | 1,864 (11.8) | 4,399 (5.5) |

**Supplementary Table 2.** Variable Means by Positive and Negative Observation Groups of the Original Model Development and Validation Datasets.

| Variable | Validation Dataset |  |  | Original Model Development Dataset |  |  |
| --- | --- | --- | --- | --- | --- | --- |
|  | Negative | Positive | All | Negative | Positive | All |
| Length of Stay (hours) | 345 | 282 | 331 | 97 | 174 | 112 |
| Vital Sign Data Count | 25 | 29 | 27 | 30 | 41 | 32 |
| Laboratory Data Count | 5 | 7 | 6 | 43 | 53 | 45 |
| Unstructured Data Count | 13 | 16 | 15 | 58 | 60 | 59 |
| Age (years) | 68 | 74 | 69 | 61 | 65 | 62 |
| Pulse (bpm) | 76 | 79 | 77 | 79 | 83 | 79 |
| Height (in) | 69 | 69 | 69 | 66 | 67 | 67 |
| Weight (lb) | 205 | 188 | 201 | 198 | 185 | 196 |
| Systolic Blood Pressure (mmHg) | 131 | 125 | 130 | 128 | 121 | 127 |
| Diastolic Blood Pressure (mmHg) | 72 | 68 | 71 | 72 | 68 | 71 |
| Oxygen Saturation (via pulse oximetry, %) | 95 | 95 | 95 | 97 | 97 | 97 |

Abbreviations: bpm, beats per minute; in, inches; L, liter; lb, pounds; min, minutes; mmHg, millimeters of mercury.

## REFERENCES

1. Lindenfeld J, Feldman AM, Saxon L, Boehmer J, Carson P, Ghali JK, et al. Effects of cardiac resynchronization therapy with or without a defibrillator on survival and hospitalizations in patients with New York Heart Association class IV heart failure. Circulation. 2007;115: 204–212.

2. Chen J, Normand S-LT, Wang Y, Krumholz HM. National and regional trends in heart failure hospitalization and mortality rates for Medicare beneficiaries, 1998-2008. JAMA. 2011;306: 1669–1678.

3. Loehr LR, Rosamond WD, Chang PP, Folsom AR, Chambless LE. Heart failure incidence and survival (from the Atherosclerosis Risk in Communities study). Am J Cardiol. 2008;101: 1016–1022.

4. Virani SS, Alonso A, Aparicio HJ, Benjamin EJ. Heart disease and stroke statistics—2021 update: a report from the American Heart Association. Circulation. 2021. Available: https://www.ahajournals.org/doi/abs/10.1161/CIR.0000000000000950

5. Ahmad T, Lund LH, Rao P, Ghosh R, Warier P, Vaccaro B, et al. Machine learning methods improve prognostication, identify clinically distinct phenotypes, and detect heterogeneity in response to therapy in a large cohort of heart failure patients. J Am Heart Assoc. 2018;7: e008081.

6. Gayat E, Arrigo M, Littnerova S, Sato N, Parenica J, Ishihara S, et al. Heart failure oral therapies at discharge are associated with better outcome in acute heart failure: a propensity-score matched study. Eur J Heart Fail. 2018;20: 345–354.

7. McDonagh TA, Metra M, Adamo M.… Guidelines for the diagnosis and treatment of acute and chronic heart failure: Developed by the Task Force for the diagnosis and treatment of acute and chronic heart…. Eur Heart J. 2021. Available: https://academic.oup.com/eurheartj/article-abstract/42/36/3599/6358045

8. van der Meer P, Gaggin HK, Dec GW. ACC/AHA Versus ESC Guidelines on Heart Failure: JACC Guideline Comparison. J Am Coll Cardiol. 2019;73: 2756–2768.

9. Gustafsson F, Rogers JG. Left ventricular assist device therapy in advanced heart failure: patient selection and outcomes. Eur J Heart Fail. 2017;19: 595–602.

10. Rossignol P, Hernandez AF, Solomon SD, Zannad F. Heart failure drug treatment. Lancet. 2019;393: 1034–1044.

11. Csepe TA, Kilic A. Advancements in mechanical circulatory support for patients in acute and chronic heart failure. J Thorac Dis. 2017;9: 4070–4083.

12. Mpanya D, Celik T, Klug E, Ntsinjana H. Machine learning and statistical methods for predicting mortality in heart failure. Heart Fail Rev. 2021;26: 545–552.

13. Angraal S, Mortazavi BJ, Gupta A, Khera R, Ahmad T, Desai NR, et al. Machine Learning Prediction of Mortality and Hospitalization in Heart Failure With Preserved Ejection Fraction. JACC Heart Fail. 2020;8: 12–21.

14. Samuel OW, Asogbon GM, Sangaiah AK, Fang P, Li G. An integrated decision support system based on ANN and Fuzzy_AHP for heart failure risk prediction. Expert Syst Appl. 2017;68: 163–172.

15. Eapen ZJ, Liang L, Fonarow GC, Heidenreich PA, Curtis LH, Peterson ED, et al. Validated, electronic health record deployable prediction models for assessing patient risk of 30-day rehospitalization and mortality in older heart failure patients. JACC Heart Fail. 2013;1: 245–251.

16. McGilvray MMO, Heaton J, Guo A, Masood MF, Cupps BP, Damiano M, et al. Electronic Health Record-Based Deep Learning Prediction of Death or Severe Decompensation in Heart Failure Patients. JACC: Heart Failure. 2022. doi:10.1016/j.jchf.2022.05.010

17. VHA Synthetic Data Lake (V-CHAMPS). Available: https://www.data.va.gov/stories/s/How-to-Access-Synthetic-Data-in-the-VA/rssm-v4rt/

18. Breiman L. Random Forests. Mach Learn. 2001;45: 5–32.

19. Raiaan MAK, Mukta MSH, Fatema K, Fahad NM, Sakib S, Mim MMJ, et al. A review on large language models: Architectures, applications, taxonomies, open issues and challenges. IEEE Access. 2024;12: 26839–26874.

20. Zhou Z, Ning X, Hong K, Fu T, Xu J, Li S, et al. A survey on efficient inference for large Language Models. arXiv [cs.CL]. 2024. Available: http://arxiv.org/abs/2404.14294

21. Wong J, Horwitz MM, Zhou L, Toh S. Using machine learning to identify health outcomes from electronic health record data. Curr Epidemiol Rep. 2018;5: 331– 342.

22. Goldstein BA, Navar AM, Pencina MJ, Ioannidis JPA. Opportunities and challenges in developing risk prediction models with electronic health records data: a systematic review. J Am Med Inform Assoc. 2017;24: 198–208.

23. Si Y, Du J, Li Z, Jiang X, Miller T, Wang F, et al. Deep representation learning of patient data from Electronic Health Records (EHR): A systematic review. J Biomed Inform. 2021;115: 103671.

24. Xiao C, Choi E, Sun J. Opportunities and challenges in developing deep learning models using electronic health records data: a systematic review. Journal of the American Medical Informatics Association. 2018;25: 1419–1428.

25. Petmezas G, Papageorgiou VE, Vassilikos V, Pagourelias E, Tsaklidis G, Katsaggelos AK, et al. Recent advancements and applications of deep learning in heart failure: Α systematic review. Comput Biol Med. 2024;176: 108557.

26. Alsaify AR, Siam A, Hassan H, Alzubaidi M, Househ M. The Use of Deep Learning in the Diagnosis and Prediction of Heart Failure: A scoping review. Proceedings of the 2024 8th International Conference on Medical and Health Informatics. New York, NY, USA: ACM; 2024. pp. 186–192.

27. Liu T, Krentz A, Lu L, Curcin V. Machine learning based prediction models for cardiovascular disease risk using electronic health records data: systematic review and meta-analysis. Eur Heart J Digit Health. 2025;6: 7–22.

